# Functional homologous recombination assay on FFPE specimens of advanced high-grade serous ovarian cancer predicts clinical outcomes

**DOI:** 10.1101/2022.10.01.22280598

**Authors:** Sanna Pikkusaari, Manuela Tumiati, Anni Virtanen, Jaana Oikkonen, Yilin Li, Fernando Perez-Villatoro, Taru Muranen, Matilda Salko, Kaisa Huhtinen, Anna Kanerva, Heidi Koskela, Johanna Tapper, Riitta Koivisto-Korander, Titta Joutsiniemi, Ulla-Maija Haltia, Heini Lassus, Sampsa Hautaniemi, Anniina Färkkilä, Johanna Hynninen, Sakari Hietanen, Olli Carpén, Liisa Kauppi

## Abstract

Deficiency in homologous recombination (HR), a key DNA repair pathway, is a defining characteristic of many high-grade serous ovarian carcinomas (HGSC) and a major determinant of therapy outcomes. Patients with HR-deficient (HRD) tumors are more sensitive to DNA damaging platinum-based chemotherapy and HR deficiency also confers sensitivity to PARP inhibitors. While PARP inhibitors are highly effective in some patients, they are expensive and not without side effects, thus it is imperative to identify patients most likely to benefit from them. We set out to develop a clinically feasible assay for identifying functionally HRD tumors based on the detection of RAD51, a key HR protein. Our functional HR assay can be performed on FFPE tumor sample sections obtained from both treatment-naïve and neoadjuvant chemotherapy treated HGSC patients. We show that the functional HR test predicts key clinical outcomes, including platinum-free survival and overall survival after PARPi treatment. Our results indicate that RAD51-based HRD testing has great potential to predict platinum and PARPi sensitivity in the clinical setting.

## Introduction

DNA-damaging agents constitute the cornerstone of many classical cancer therapies. For tubo-ovarian high-grade serous carcinoma (HGSC), the most common and most lethal gynecologic malignancy, platinum derivatives are used as DNA-damaging compounds in standard-of-care. Platinum intercalates with DNA, causing inter- and intrastrand links, which result in single- and double-stranded DNA breaks in S phase of cell cycle, while DNA is replicated. These lesions are lethal to the cell if not repaired. The key DNA repair pathway for overcoming platinum-induced DNA damage is homologous recombination repair (HRR). Accordingly, HGSCs with high HR capacity are typically platinum-resistant, while HR-deficient (HRD) ones are at least partially platinum-sensitive. Moreover, HRD tumors are also sensitive to PARP inhibitors (PARPi, Bryant et al., 2005; Farmer et al., 2005). PARPi maintenance therapy has transformed the HGSC management in the last few years (Banerjee et al., 2020; Mirza et al., 2016). The outcomes of PARPi therapy strongly correlate with platinum sensitivity (Coleman et al., 2017; del Campo et al., 2019; Ledermann et al., 2016; Swisher et al., 2021b).

Given the markedly better response of patients with HRD tumors to both platinum and PARPi, they ideally should be identified already at the time of diagnosis, that is, prior to starting drug treatment. Conversely, patients with HR-proficient tumors could be channeled to alternative drug regimens. Since the advent of PARPi in clinical use, HRD testing has been a topic of high interest, and several approaches exist to identify HRD tumors (reviewed in Miller et al., 2020; van Wijk et al., 2022). The two main ones include *(1)* sequencing of key DNA repair genes, such as *BRCA1, BRCA2* or *PALB2*, to identify pathogenic variants, both germline and somatic (e.g. Pennington et al., 2014), *(2)* quantifying HRD-associated genomic features in tumor DNA (e.g. Alexandrov et al., 2013; Davies et al., 2017; Telli et al., 2016), with often these two combined (Mirza et al., 2016; Stover et al., 2020). In addition, *ex vivo* functional assays of DNA damage induction and repair have been developed (Graeser et al., 2010; Meijer et al., 2018; Mukhopadhyay et al., 2012; Naipal et al., 2014; Tumiati et al., 2018; van Wijk et al., 2020). Recently, detection of RAD51, a central HR protein, in formalin-fixed paraffin-embedded (FFPE) sections without externally induced DNA damage has shown great promise in identifying HRD breast cancers (Castroviejo-Bermejo et al., 2018; Cruz et al., 2018; Pellegrino et al., 2022). A similar approach has recently been employed also to gynecological cancers, including some cases of advanced HGSC (Hoppe et al., 2021; van Wijk et al., 2021).

In the clinic, HRD stratification currently rests on HR repair gene mutation testing and HRD-associated genomic features (e.g. Myriad Genome Instability Score, GIS). These genomics-based HRD tests, however, have limitations. HRD is often, but not always associated with inactivating mutations in DNA repair genes, and the functional consequences of many mutations are difficult to infer (Plon et al., 2008). Genomic scars and HRD-associated mutational signatures report on “historical”, but not necessarily current HRD status of the tumor. This poses a problem because restoration of HR capacity *in vivo* occurs at appreciable frequency (Norquist et al., 2011), a process that will be missed if relying on HRD genomic signatures. Furthermore, genomic scar-based HRD estimates can be determined reliably from a specimen only when tumor percentage is higher than 30% (Myriad myChoice). Such tumor percentage is rarely attained for many HGSC patients treated with neoadjuvant chemotherapy (NACT), who constitute a substantial proportion of all cases (>45%, Knisely et al., 2020).

With PARPi having entered wide-spread clinical use as frontline maintenance therapy, there is an urgent unmet need for new and improved methods for accurately and comprehensively identifying those patients who can benefit from this potent but rather costly treatment (Wethington et al., 2021). Performing functional HRD testing on FFPE specimens is an attractive concept, since such samples are taken routinely from each patient for histopathological analysis and accordingly, this approach was named a priority by the ESMO Translational Research and Precision Medicine Working Group (Miller et al., 2020). Its utility for predicting real-world therapy sensitivity in advanced HGSC remains to be validated. Importantly, thus far HRD has not been systematically quantified from specimens obtained at interval debulking surgery (IDS), i.e., from surgery after NACT.

The herein presented RAD51-based functional HR (fHR) test of FFPE sections quantifies the real-time function of HR repair in the tumor, does not require high tumor cell content and works on both treatment-naive and NACT-treated HGSC specimens. Thus, it can overcome many of the limitations the genomics-based methods have. We show that the fHR test predicts platinum-free interval and other key clinical outcomes in HGSC patients, including overall survival after PARPi treatment in the recurrent setting. The fHR test provides a valuable tool to support clinical decision-making for platinum or PARPi treatment.

## Materials and Methods

### Patients

Fresh tumor tissue specimens were collected from consenting patients who underwent surgery for advanced HGSC at Turku University Hospital or Helsinki University Hospital. All patients were diagnosed with FIGO stage IIIB or higher HGSC. Frontline chemotherapy consisted of platinum/taxane combination. The study was conducted in accordance with the ethical principles of WMA Declaration of Helsinki and approved by the ethics boards of Hospital District of Southwest Finland and Hospital District of Helsinki and Uusimaa.

117 samples collected from 74 patients in Turku University Hospital were included in the study. These included both NACT-treated patients (n=33), undergoing diagnostic laparoscopy and/or interval debulking surgery (IDS), and patients undergoing primary debulking surgery (PDS, n=41). Thirty-five of these patients received bevacizumab during chemotherapy and as maintenance treatment. All patients had at least one year of follow-up time from diagnosis. These samples comprised the discovery and validation cohorts of chemo-naive samples, as well as the discovery cohort of NACT-treated samples (**Table 1**.).

**Table 1.**
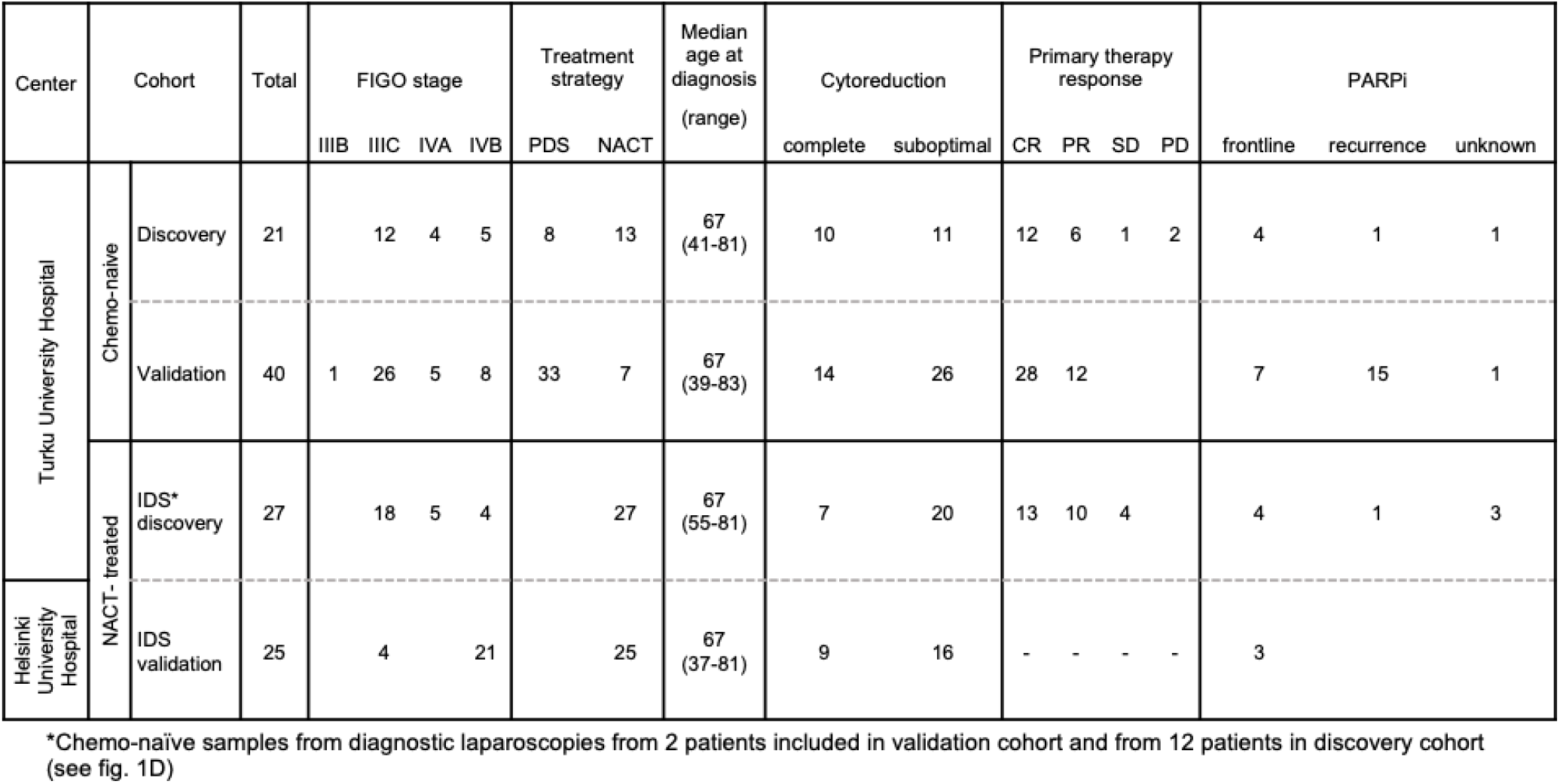
Clinicopathological characteristics of patients included in the cohorts. Complete cytoreduction refers to no residual tumor, suboptimal to >0 mm. Abbreviations: PDS, primary debulking surgery; NACT, neoadjuvant chemotherapy, CR, complete response; PR, partial response; SD, stable disease; PD, progressive disease.

For the validation cohort of NACT-treated samples additional 25 samples collected from 25 patients in Helsinki University Hospital were included in the study (**Table 1**.). Samples from these patients were available from IDS surgeries only. Bevacizumab was used in the treatment of 20 patients and all patients had at least six months of follow-up time from diagnosis.

### Chemo-naive sample cohort

The chemo-naive samples were collected at Turku and consisted of samples from 61 patients further divided into discovery and validation cohorts. The discovery cohort consisted of 21 patients and a total of 31 chemo-naive samples (**Table 1**). Eight patients belonged to the PDS treatment arm and 13 to the NACT-treatment arm. Four patients received PARPi treatment as frontline maintenance therapy, one patient at recurrence and PARPi treatment status of one (PARPi trial) patient was unknown. The validation cohort consisted of 40 unselected patients and a total of 59 chemo-naive samples (**Table 1**). Thirty-three patients belonged to the PDS treatment arm and seven to the NACT treatment arm. Seven patients received PARPi as frontline maintenance therapy, 15 patients at recurrence and PARPi treatment status of one (PARPi trial) patient was unknown.

### IDS sample cohort (NACT-treated)

NACT-treated samples were obtained from interval debulking surgery from 27 patients at Turku University Hospital (**Table 1**.). Samples obtained from diagnostic laparoscopy from 12 of these patients were included also in the discovery cohort and 2 in the validation cohort of chemo-naive samples.

Thirteen patients had only IDS samples available. Four patients received PARPi as frontline maintenance therapy, one patient at recurrence and the PARPi treatment status of three (PARPi trial) patients was unknown. All patients had received 3 cycles of chemotherapy prior to IDS, except for one, who had received 5 cycles. Time from last NACT infusion to IDS was on average 30 days (range 15-40 days). Additional NACT-treated samples from 25 patients were obtained from IDS at Helsinki (**Table 1**.) Three of these patients received PARPi treatment as frontline therapy.

### PARPi frontline-treated patients

Of the patients in the above cohorts, 15 received PARPi as frontline maintenance treatment. Five of these patients were estimated to have lost either BRCA1 or BRCA2 function in their tumor as a consequence of somatic or inherited mutations, and a subsequent somatic loss-of heterozygosity (referred to as BRCAmut throughout, see **STable 1**. for details). Seven were assumed BRCA wild type since no mutations were detected in the whole genome sequencing (WGS). Additionally, three patients, of which only clinical BRCA1/2 genetic testing results were available, were found to be BRCAmut (one patient with non-pathogenic variant, **STable 1**.).

### PARPi-at-recurrence patients

Of the patients in the above cohorts, 17 received PARPi treatment at recurrence. Five of these patients were BRCAmut and 12 patients were BRCA wild type, as estimated from WGS.

### Tumor material for FFPE blocks

Tumor specimens for FFPE blocks were fixed in formalin within two hours after surgical removal. The short time-to-fixation was found to be critical; samples with time-to-fixation longer than four hours often displayed notable tissue degeneration, as assessed from histological slides by a trained pathologist and the amount of DNA damage, marked by γH2Ax, varied considerably compared to a sample with less than two hours to fixation (**SFig. 1**). Samples with time-to-fixation longer than four hours were not included in this study.

The following anatomical tumor locations were sampled: ovary, adnexa and/or fallopian tube, collectively referred to as tubo-ovarian, as well as omentum and peritoneum. Twenty-one patients had samples available from more than one anatomical location.

### Relapse samples

During relapses, samples were collected from ascites or surgery. Relapse samples were available for 17 patients, of whom 11 had ascites, 1 pleural fluid and 7 surgical samples from peritoneum, bowel, lymph node or mesenterium.

### Histology and immunofluorescence stainings

Four μm-thick FFPE sections of tumor specimens were deparaffinized in xylene and rehydrated in decreasing concentrations of ethanol. Sections were boiled in 10 mM citrate buffer, pH 6, for 20 min for antigen retrieval and immunostained according to standard procedures with primary antibodies against cytokeratin 7 (CK7, diluted 1:500, ab9021 Abcam), γH2Ax (diluted 1:1000, ab11174 Abcam), geminin (diluted 1:200, ab104306 Abcam) and RAD51 (diluted 1:1000, ab133534 Abcam) in three combinations: CK7-γH2Ax, geminin-γH2Ax and geminin-RAD51. Sections were then incubated with fluorescently labeled secondary antibodies (diluted 1:500, donkey anti-mouse IgG-AlexaFluor 488, A21202; and donkey anti-rabbit IgG-AlexaFluor 647, A31573; Life Technologies) and nuclei were counterstained with Hoechst. Slides were digitized using a Pannoramic 250 FLASH II slide scanner (3DHISTECH) at 20x magnification.

### Immuno-quantification and HR scoring

For each sample, 2-3 images of 3.8 mm^2^ area in size with high tumor cell content, marked by CK7 (Chu et al., 2000; Zhang et al., 2008), and visible DNA damage, marked by γH2Ax, were chosen for analysis based on the CK7-γH2Ax stained section. The images (.tiff) were analyzed in ImageJ using custom macros. In geminin-γH2Ax and geminin-RAD51 stained sections, areas with epithelial cells were identified with the help of CK7-γH2Ax stained serial section. Next, epithelial cell nuclei were identified based on their size and shape and a mask of the nuclei was created. As homologous recombination can only be performed during the S/G2 phase of the cell cycle, we identified epithelial nuclei positive for the S/G2 phase marker geminin by applying the epithelial nuclei mask to geminin channel. Finally, we applied the geminin-positive epithelial nuclei masks to the γH2Ax and RAD51 channels, to quantify DNA damage and HR-mediated repair, respectively (see **SFig. 2** for details). To identify RAD51 and γH2Ax positive nuclei, a threshold and size limit was used followed by dilation to merge all particles inside a single nucleus together, as individual foci cannot be reliably distinguished in 20x images. Particles were then automatically counted, resulting in the total number of geminin positive nuclei, and the number of geminin positive RAD51 or γH2Ax positive nuclei per each image. From these analyses, we obtained the following quantifications per each sample: percent of S/G2 phase nuclei with DNA damage (% of γH2Ax+ and geminin+ tumor nuclei out of all geminin+ tumor nuclei) and percent of S/G2 phase nuclei performing HR-mediated repair i.e., fHR score (% of RAD51+ and geminin+ tumor nuclei out of all geminin+ tumor nuclei). For samples of which geminin-γH2Ax double-staining was not available, geminin-RAD51 and CK7-γH2Ax stained serial sections were used to estimate the amount of DNA damage in S/G2 phase cells. For chemo-naïve specimens, only samples with >10% of S/G2 phase cells with DNA damage were used to generate fHR scores; 97% (87 out of 90) of chemo-naïve specimens fulfilled these criteria. For IDS specimens, only samples with >30% of S phase cells with DNA damage were used; 98% (51 out of 52) of IDS specimens fulfilled these criteria. These cut-offs were established as the fHR score cannot be higher than the percent of S/G2 phase cells with DNA damage, so for a sample to be scored as fHRP (later determined as fHR score of ≥10 and ≥30 for chemo-naïve and IDS samples respectively) there must be at least that much of S/G2 phase DNA damage. For each sample, a minimum of 50 geminin-positive cells were analyzed. Immunofluorescence quantification of all specimens was performed blinded to BRCA mutation status and to data on clinical outcomes. Every .tiff image was analyzed 2-3 times, depending on the consistency of the result.

### Clinical parameters

Once fHR scores were determined for each patient, we analyzed them for correlations with clinical outcomes. When an fHR score was available from multiple anatomical locations from the same patient, the highest score was chosen for analyzing correlations with clinical outcomes. Patient’s response to primary therapy was determined with RECIST 1.1 criteria (Eisenhauer et al., 2009). Platinum-free interval (PFI) was calculated from the last platinum dosage of primary chemotherapy to the first disease recurrence. Since PARPi maintenance treatment in the frontline setting prolongs PFI (reviewed e.g. in Banerjee et al., 2020), for analyses with PFI as a variable, patients who received PARPi treatment as frontline maintenance therapy were excluded. For analyses with overall survival (OS) as a variable, also patients who received PARPi therapy at recurrence were excluded. Clinical trial patients whose PARPi treatment status was unknown were excluded from both OS and PFI analyses. Response to platinum therapy in the recurrent setting was defined as <6 months versus >6 months, as calculated from the beginning of the treatment to next disease progression (Eisenhauer et al., 2009; Rustin et al., 2011).

### DNA sequencing, HRR gene mutations, SBS3, ID6

Fresh frozen samples were whole-genome sequenced (WGS) and processed as described previously (Lahtinen et al., 2022). Additional WGS samples were sequenced using NovaSeq 6000 by Novogene (Novogene (UK) Co. Ltd), and further whole-exome sequencing (WES) samples using HiSeq 2000 by BGI (BGI Europe A/S, Denmark) with Agilent SureSelect human all exon V5. Samples for sequencing were only collected in Turku University Hospital. Out of 74 patients, 69 were analyzed with WGS with blood-derived matched normal and one without while four were analyzed with WES. For two of the patients with WGS, only germline analysis was possible due to lack of tumor purity.

Somatic variants were called using GATK (McKenna et al., 2010) Mutect2. Germline variants were called from WGS normals using GATK with allele-specific filtering and joint-genotyping. Germline variant allele frequencies in tumor samples were obtained with Mutect2 forced calling. Copy-number segmentation was performed using GATK and used as input in allele-specific copy-numbers and tumor purity estimation with ASCAT (Van Loo et al., 2010).

The variant data was queried for deleterious mutations in the following HR-pathway genes: BRCA1/2, BARD1, BRIP1, CHEK2, MRE11A, NBN, PALB2, RAD50, RAD51C/D, CDK12, ATM, and the FANCA/B/C/D2/E/F/G/I/L/M. The mutations were considered as deleterious if causing premature stop, frameshift, or altered splicing, or if classified as pathogenic/likely pathogenic in ClinVar (Landrum et al., 2018) release 2022-05-28. The likelihood of mutation homogeneity in tumor or loss of heterozygosity of a germline variant was estimated according to the mutation allele frequency, locus copy number, and tumor fraction in the sequenced sample.

Signatures were independently fitted for each sample using an R implementation based on SigProfilerAttribution (Alexandrov et al., 2020) against COSMIC reference signatures v3.2 (Tate et al., 2019). Indel signatures were only analyzed in WGS patients with matched normal. Single base substitutions (SBS) signatures were adjusted for trinucleotide frequency of GRCh38 (WGS) or Agilent V5 targets (WES) without chromosomes Y and M. Signatures with at least 20% ovarian cancer occurrence in COSMIC were used as starting signatures. SBS60 was also included for WES samples.

Samples were considered HRD for SBS3 if the signature contribution was greater than zero while for ID6, a threshold of 0.2796 was used. Unit length normalized indel spectra were clustered with 2-means and the threshold that maximized F1-score predicting assignment to the high microhomology deletion cluster was chosen. Each sample’s signature-derived HRD status was selected from a sequenced sample with matching tissue whenever possible. A minimum purity of 5% was required for HRD and 10% for HRP. SBS3 was estimated for 83 samples from 71 patients and ID6 based status was available for 77 samples from 67 patients.

### Genomic homologous recombination deficiency test (ovaHRDScar)

HR deficiency causes characteristic loss-of-heterozygosity (LOH), large scale transitions (LSTs) and telomeric allelic imbalances (TAI) that can be quantified and used to identify HRD tumors (Takaya et al., 2020; Telli et al., 2016). A recently optimized algorithm (called ovaHRDScar), to quantify these allelic imbalances in HGSC samples, was used for the classification of the samples as HRD or HRP (Perez-Villatoro et al., 2021). A cut-off value of >54 for the sum of LOH, LSTs and TAI was used for classifying a sample as HRD.

Tumor purity of the samples was estimated based on 1) somatic copy-number profiles using the software ASCAT, 2) variant allele frequency of the truncal mutation in gene TP53 (TP53-VAF), using the formula: 2 / ((CN / TP53-VAF) - (CN - 2)), where CN corresponds to the absolute copy-number value estimated by ASCAT in the corresponding truncal mutation locus, and/or 3) visual estimation from H&E sections by a trained pathologist. The highest purity value resulting from the three criteria was selected. Samples with purity below 25% were ignored.

### Statistical analyses

Statistical analyses were performed with GraphPad Prism 9.0 software. A p-value of <0.05 was considered significant. The chosen statistical test for each analysis is indicated in figure legends.

## Results

### Functional HR (fHR) status

The fHR test established here is similar to functional, RAD51-based HR assays described before (Castroviejo-Bermejo et al., 2018; Cruz et al., 2018; Hoppe et al., 2021; Pellegrino et al., 2022; van Wijk et al., 2021). One key difference is that we score S/G2 phase nuclei as RAD51-positive or - negative whereas others quantified RAD51-positive nuclei based on individual RAD51 focus counts (≥5 foci per nucleus, Pellegrino et al., 2022; ≥2 foci per nucleus van Wijk et al., 2021). Thus, fHR scoring can be performed from digitized slides that are scanned at a low (20x) magnification. In contrast to earlier work, specimens analyzed were all advanced HGSCs. Importantly, the fHR protocol was employed and optimized, for the first time, to NACT-treated (IDS) samples as well (**Fig. 1A**). Chemo-naive samples as well as NACT-treated samples were initially examined in discovery cohorts, and subsequently in validation cohorts (**Table 1**.).

**Figure 1.**
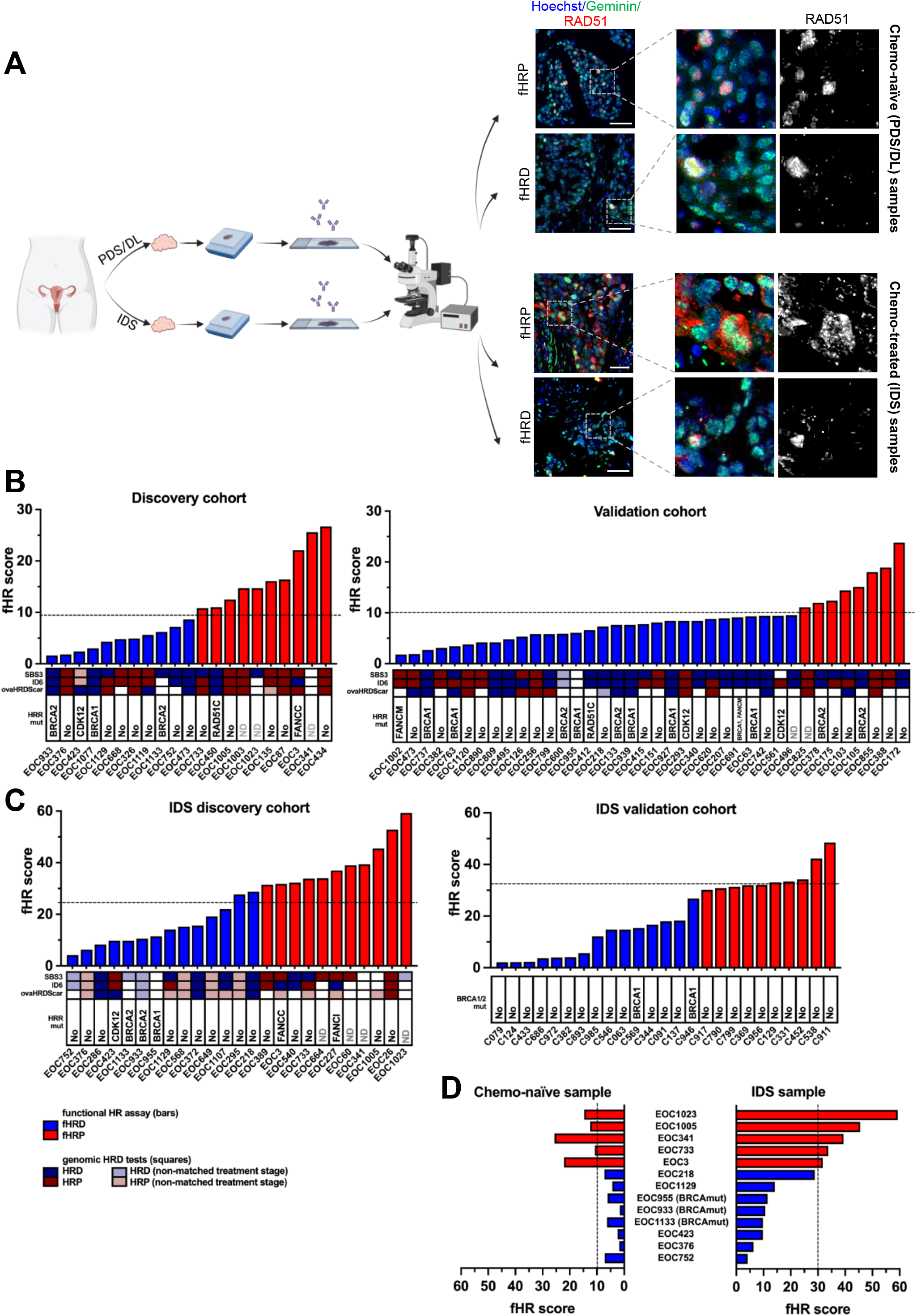
**A**. Workflow for functional HR assay with fHRD and fHRP example images of geminin-RAD51 double stained chemo-naive samples and NACT-treated (IDS) samples. **B-C**. Distribution of fHR scores in the (B) discovery and validation cohorts of chemo-naïve samples, as well in the (C) NACT-treated (IDS) discovery and validation cohorts. The highest fHR score per patient was chosen if a score was available from >1 sample. Dashed line indicates proposed fHRD vs fHRP cut-offs. The results from available genomics-based HRD assays are shown in squares, blue shades indicating HRD and red HRP (non-matched treatment stage refers to a sequencing sample that was obtained at a different surgery (PDS/DL vs IDS) than the sample for fHR analysis). Deleterious mutations in HRR genes identified from WGS/WES data are indicated for each patient (ND = no data). For IDS validation cohort, only BRCA1/2 mutational testing result from the clinic was available. **D**. Comparison between the fHR score of patient-matched chemo-naive and NACT-treated (IDS) samples.

We were able to obtain a fHR score from 94% of samples (134**/**143 samples). Of the nine samples for which a fHR score could not be determined, three were excluded because the amount of DNA damage in S/G2 phase cells did not exceed the set thresholds (>10% for chemo-naive, >30% for NACT-treated samples), three because they did not have enough geminin positive tumor cells (threshold of >50 cells), and another three due to failed staining or imaging. Because of these unanalyzable samples, we had to exclude two patients from the validation cohort (n=40->38) and two from the IDS cohort (n=27->25) from analyses with fHR score.

### Defining fHRD versus fHRP categories for chemonaive specimens

In chemonaive specimens from patients in the discovery cohort (n= 21), fHR scores ranged from 1.6 to 26.7. Scores were bimodally distributed, consistent with two HR categories (**SFig 3**). The cut-off value of 10% RAD51+ geminin+ cells (that is, fHR score 10.0) was chosen to define fHRD versus fHRP in chemonaive specimens; this value was recently determined to be highly predictive of HRD in a cohort of >100 breast cancer patients, as assessed by PDX-based PARPi sensitivity (Pellegrino et al., 2022). Using this cut-off, half of discovery cohort patients – and all three BRCAmut cases therein – fell into the fHRD category (**Fig. 1B**), as expected (Konstantinopoulos et al., 2015).

In the validation cohort (n=38) fHR scores ranged from 1.8 to 23.8. When applying the 10% cut-off value for fHRD, 79% (n=30) of the patients fell into the fHRD category and 21% (n=8) into the fHRP category (**Fig 1B**). The validation cohort included 11 BRCAmut patients, nine of which were categorized as fHRD and two as fHRP. The large proportion of BRCAmut patients (29%) explains, at least in part, the higher proportion of fHRD patients in the validation cohort compared to the discovery cohort (14% BRCAmut).

We employed hazard ratio analysis to further test how the proposed fHRD cut-off value (10%), as opposed to other cut-off values, e.g. 15% used by van Wijk et al. (2021), correlates with PFI. Setting the cut-off to 15 or 9 also produced significant results, although the 95% confidence interval was narrower with the cut-off of 10 (**SFig. 4**).

For 21 patients, chemo-naive samples from more than one anatomical location were analyzed. While numerical values of fHR scores varied between the locations, the fHR category (fHRP or fHRD) remained the same in the different locations sampled (**SFig. 5**) in all cases when the 10% cut-off was employed.

### Defining fHRD versus fHRP categories for IDS specimens

Half or more of HGSC patients undergo NACT, followed by IDS, and chemonaive samples are typically not available for these patients. Thus, it is critical to also be able to estimate fHR in samples obtained at IDS. We set out to quantify fHR for this type of specimens in an IDS discovery cohort of 27 NACT-treated patients. Since no precedent of RAD51 scoring in IDS specimens exists in the literature, we first examined the features of RAD51 staining (**Fig. 1A**) as well as numerical values of fHR scores in these samples. Compared to chemo-naive samples, the overall staining pattern was less clean and cytoplasmic RAD51 signal was observed more often (**Fig. 1A** and **SFig. 6**). For fHR quantification, only nuclear RAD51 signal was considered. In the discovery cohort of IDS samples, fHR scores were substantially higher than in chemonaive samples, ranging from 4.2 to 59.3, and the three BRCAmut patients in this cohort had fHR values of 9.8, 10.6 and 11.5 (**Fig. 1C**). Taken together, these findings indicate that the appropriate cut-off value for fHRD in IDS specimens should be set higher than the 10% defined for chemo-naive specimens.

To estimate a cut-off value for fHRD versus fHRP in IDS specimens, we examined their fHR score distribution. As with chemonaive samples, fHR values followed a bimodal distribution (**SFig. 3**). Setting the cut-off value for fHRD at <30% RAD51+ gem+ cells, 54% of our IDS discovery cohort patients (14/25) fell into the fHRD category, in line with the estimated percentage of HRD tumors in HGSC (Konstantinopoulos et al., 2015) and TNBC (Pellegrino et al., 2022). We note that when using the 30% cut-off value for IDS samples, the last patient with SBS3+, ID6+ and ovaHRDScar+ features (EOC218) is captured as fHRD, as are all three BRCAmut patients (**Fig. 1C**).

In the IDS validation cohort (n=25), fHR scores ranged from 2.1 to 48.5. When applying the 30% cut-off value, 40% (n=10) of the patients fell into the fHRP category and 60% (n=15) into the fHRD category (**Fig. 1C**). The two BRCAmut patients had fHR scores of 15.4 and 26.8 and fell in the fHRD category.

To further test how the proposed fHRD cut-off value (30%) correlates with PFI, we employed hazard ratio analysis. A cut-off of 25 and 35 also produced a significant result, but the 95% confidence interval was narrower with the cut-off of 30 (**SFig. 4)**.

### fHR status and HRR gene mutations

Given that biallelic deleterious mutations in HRR genes should impair functional HR, one would expect that tumors with such mutations are fHRD. In this context, BRCAmut status is generally considered the ground truth for HRD. Altogether, our study included 18 patients with BRCAmut tumors (out of a total n=99 individual patients, chemo-naive and IDS cohorts considered jointly), but two of these were unscorable, due to insufficient DNA damage and tumor cells. Fourteen out of 16 BRCAmut tumors (88%) were scored as fHRD. The two fBRCAmut samples scored as fHRP were tumors with somatic *BRCA2* mutations, and both were found to have reading frame restoring mutations in their relapse samples that were analyzed by WGS (**STable 1**.).

Two patients (EOC450 and EOC412) carried a deleterious mutation in *RAD51C*, a well-established HRR gene (**STable 1**.). EOC412 was scored as fHRD (fHR score = 6,6), but EOC450 was scored as fHRP, when using the 10% cut-off (fHR score = 11.0, **Fig. 1B**). This is not surprising, given that RAD51C depletion reduces but does not abolish RAD51 recruitment to sites of DNA damage (Chun et al., 2013; Han et al., 2017). This example illustrates a limitation of the RAD51-based fHR assay: it cannot identify all functionally HRD samples, only those with impaired RAD51 loading.

The three *CDK12*mut tumors were scored as fHRD (**Fig. 1B**). Samples with biallelic loss of *FANCC* or *FANCI* were scored as fHRP, whereas the one patient with intact BRCA1/2 and biallelic loss of FANCM, due to germline c.5101C>T, pQ1701X and loss-of-heterozygosity, was fHRD. (**Fig. 1B and 1C**).

### fHR and SBS3, ID6 and ovaHRDscar status

For most of the patients sampled at Turku University Hospital, we had, in addition to the fHR score, information available from at least one of the following genomics-based HRD estimates: mutational signature SBS3, mutational signature ID6 (Alexandrov et al., 2013) and/or ovaHRDScar (Perez-Villatoro et al., 2021). For 46% of the patients (n=34), at least one of the available methods for HRD determination disagreed with the other estimates (**Fig. 1B** and **1C**). In addition, patients without genomic HRD-associated features were found in the fHRD category and vice versa.

### Comparison between paired chemo-naive and IDS samples

For 13 patients, we obtained fHR scores from both pre- and post-NACT specimens (from diagnostic laparoscopy and IDS), allowing us to assess whether fHR status determined from these two sample types agreed with each other. We found that pre- and post-NACT samples from all 13 patients fell into the same fHR category **(Fig. 1D**). Although numerical fHR scores are higher in IDS samples, NACT treatment does not seem to impact fHR status (fHRD versus fHRP), as classified using our threshold.

### Low fHR scores correlate with better primary therapy response

Platinum derivatives, used in primary therapy of HGSC, induce DSBs and thus are expected to result in better primary therapy response in patients with HRD tumors. Primary therapy response assessment was available for patients sampled at Turku University Hospital. We correlated fHR scores with primary therapy response and found that low fHR scores, indicative of HR deficiency, were enriched in complete and partial response groups. The median of fHR scores was significantly lower in patients with a complete response, compared to patients with stable or progressive disease in both chemo-naïve (6.6 vs. 16.4) and NACT-treated (12.8 vs. 45.9) samples. Also, in both groups, the median fHR score was significantly lower in the partial response group than in the stable or progressive disease group (9.1 vs. 16.4 and 27.7 vs. 45.9) (**Fig. 2A**).

**Figure 2.**
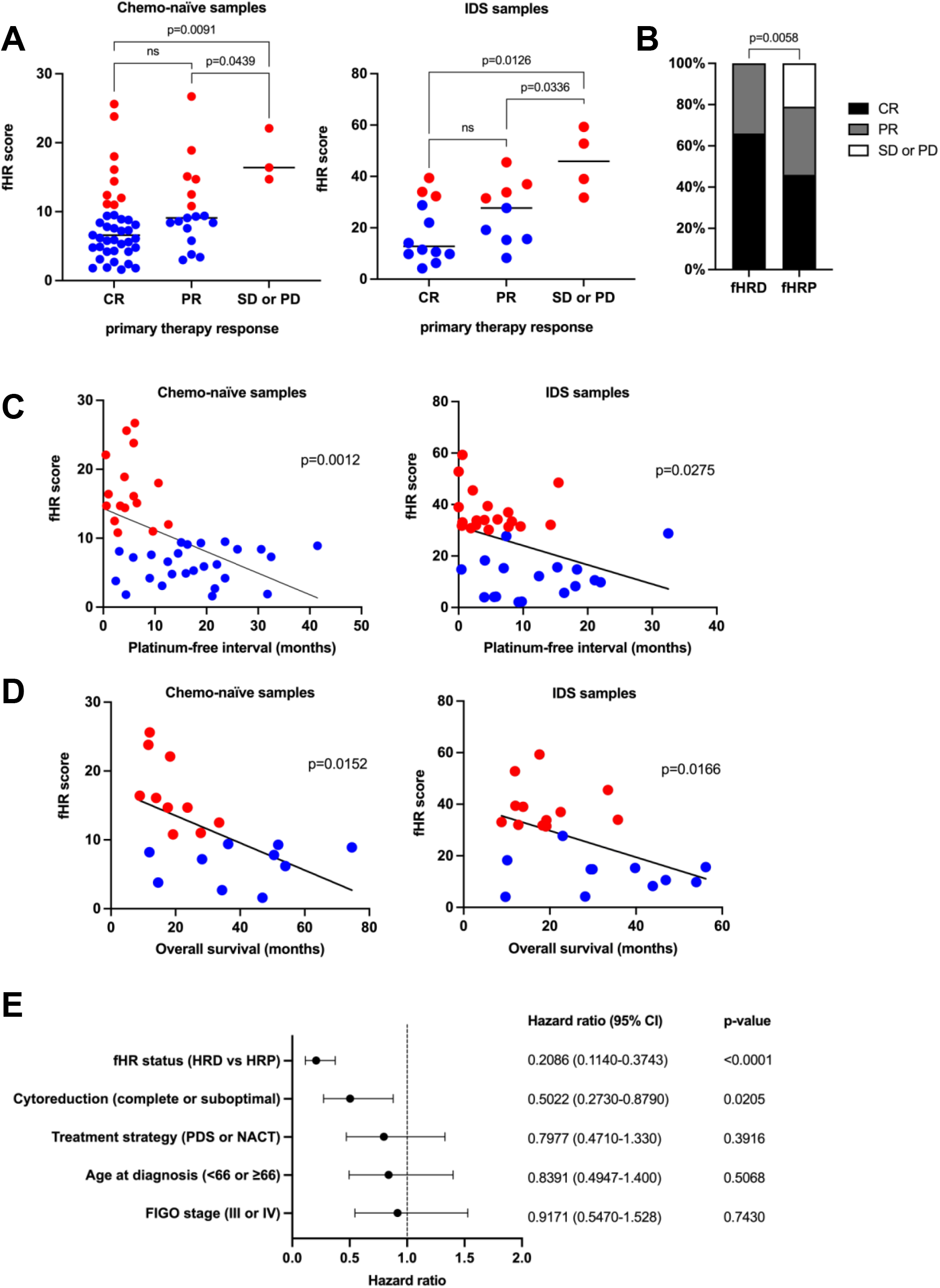
Low fHR scores correlate with longer PFI. **A**. Median of fHR scores is lower in CR and PR groups compared to SD/PD (Mann-Whitney test). **B**. Patients classified as fHRP have SD/PD after primary therapy more often than fHRD patients (Fisher’s exact). **C-D**. PFI and OS significantly correlate with lower fHR scores (linear regression). **E**. Comparison of the fHR classification with other prognostic clinical parameters. The fHRD status and success of cytoreduction were found to significantly correlate with longer PFI (multivariate Cox proportional hazards regression).

We then divided patients into fHRD and fHRP categories, based on their tumor’s fHR score, using the cut-off values determined above. In the fHRD category, 66% of the patients had complete response to primary therapy and the rest (34%) had partial response. In the fHRP category, 46% of patients had complete response, 33% partial response and 21% of patients had stable or progressive disease. The proportions of primary therapy response categories were significantly different in fHRD and fHRP categories (**Fig. 2B**). Primary HGSC therapy consists of surgery and platinum therapy, but surgical results did not differ between fHRD and fHRP groups (**SFig.7**), thus the better primary therapy response in the fHRD group is due to better response to platinum-based chemotherapy.

### Low fHR scores correlate with longer PFI and OS

Next, we assessed whether the numerical fHR score correlated with platinum-free interval (PFI) and overall survival (OS) of the patients. The fHR score, obtained from both chemo-naïve and NACT-treated samples, was negatively correlated with both PFI and OS (**fig. 2C, D**).

We performed a multivariate hazard ratio analysis with fHR status and clinical parameters, namely treatment strategy (PDS vs NACT), success of cytoreduction (optimal vs suboptimal), age at diagnosis (<66 vs ≥66) and FIGO stage (III vs IV). Only fHR status (hazard ratio: 0.1982) and success of cytoreduction (hazard ratio: 0.5033) were found to significantly correlate with longer PFI (**Fig. 2E**).

### Functional HRD status correlates with platinum-free interval and overall survival

To investigate further the clinical significance of fHR status, we compared the PFI of fHRD and fHRP groups in Kaplan-Meier survival analysis. The PFI-based survival curves were significantly different between the fHRD group and the fHRP group in both chemo-naive and IDS cohorts (**Fig. 3A**). Survival curves are shown separately for the discovery and validation cohorts in **SFig. 8A**. In the chemo-naive cohort, median PFI in the fHRD group was 18.9 months, while in the fHRP group it was only 4.5 months. In the IDS cohort, two patients in the fHRP group progressed during primary therapy; such cases are excluded from the survival curves. Without these two patients, the median PFI in the IDS cohort was 15.3 months in the fHRD group and 4.5 months in the fHRP group. Patients classified as fHRD had better response to platinum-based chemotherapy as second-line treatment also (**SFig. 8B**).

**Figure 3.**
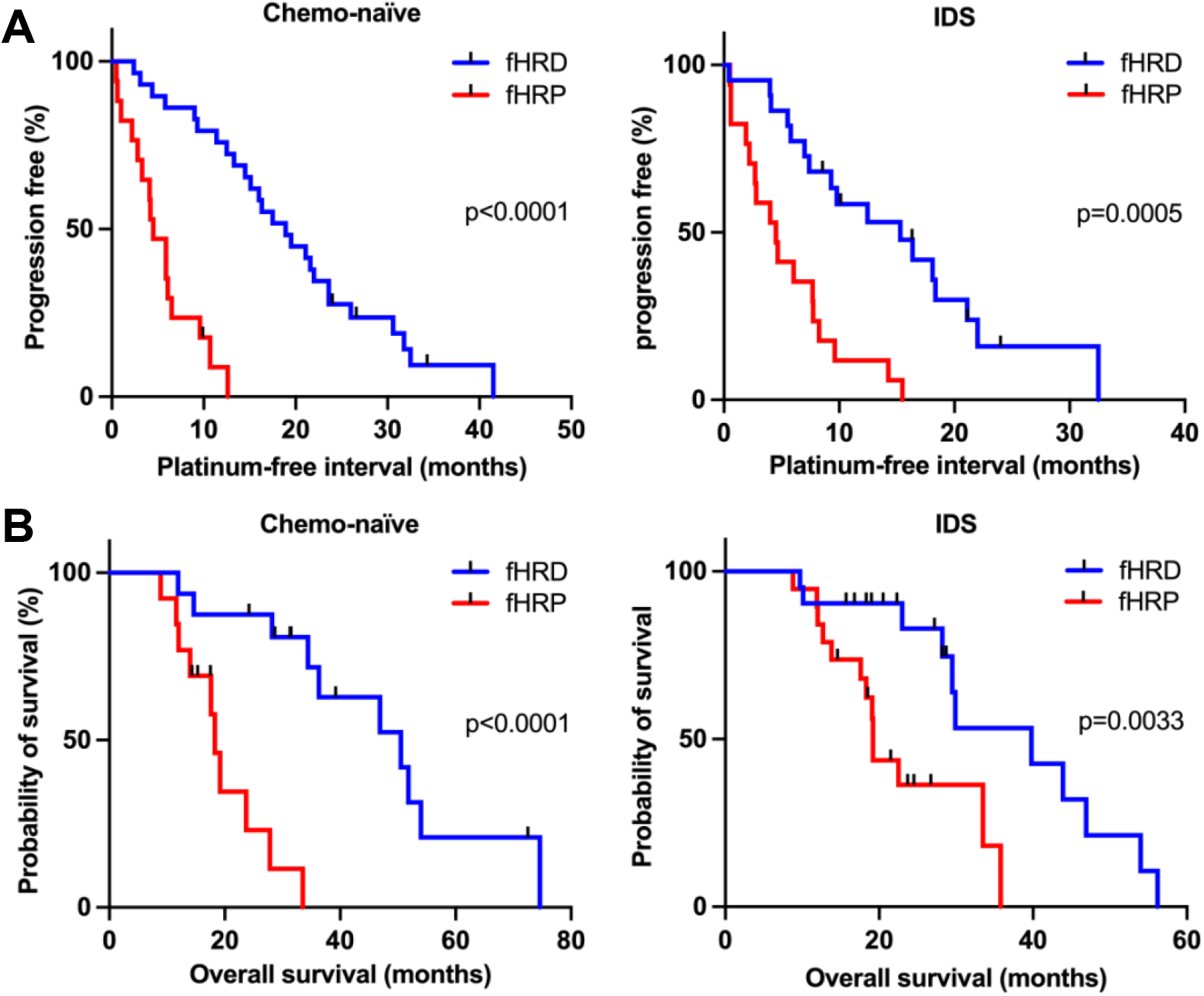
Kaplan-Meier survival analysis. **A**. fHRD status significantly predicts longer PFI and **B**. OS in both chemo-naïve and IDS cohorts (Log-rank, Mantel-Cox test).

To determine if the difference in PFI between fHRD and fHRP groups was driven primarily by the BRCAmut cases in the fHRD group, we ran the survival analysis excluding these patients. PFI-based survival curves were still significantly different between the fHRD BRCA-wildtype group (BRCAwt) and the fHRP group in both chemo-naive and IDS cohorts (**SFig. 8C**).

In the OS analysis, survival curves were significantly different between the fHRD and fHRP groups in both chemo-naive and IDS cohorts (**Fig. 3B**). In the chemo-naive cohort, median OS was 50.5 months in the fHRD group and 18.3 months in the fHRP group. In the IDS cohort, median OS for fHRD and fHRP was 39.8 months and 19.2 months, respectively.

### Functional HRD status associates with PARPi response

In addition to predicting platinum response, and at least as pertinent given the current HGSC therapy options, is to predict PARPi response. Of the patients analyzed here for fHR status, a subset had received PARPi treatment either in the frontline setting (n=15) or at recurrence (n=17, **table 1**). Although patient numbers are small and PARPi regimens are heterogeneous, these real-world data allowed for *post hoc* assessment of how fHR status associates with *in vivo* PARPi response. Because only two patients in the frontline PARPi-treated cohort were fHRP, we were unable to perform any survival comparisons in this cohort.

In the PARPi-at-recurrence cohort, where four patients were fHRP and 13 patients were fHRD, we compared the time from the start of PARPi treatment to disease progression between fHRD and fHRP groups. Although survival curves separated in favor of the fHRD group, the difference between fHRD and fHRP groups did not reach statistical significance (**Fig. 4A**). Notably, the fHR test was able to identify as fHRD those PARPi-at-recurrence -treated patients with longer OS (**Fig. 4B**), although all these patients were platinum sensitive (criterion to be eligible for PARPi treatment).

**Figure 4.**
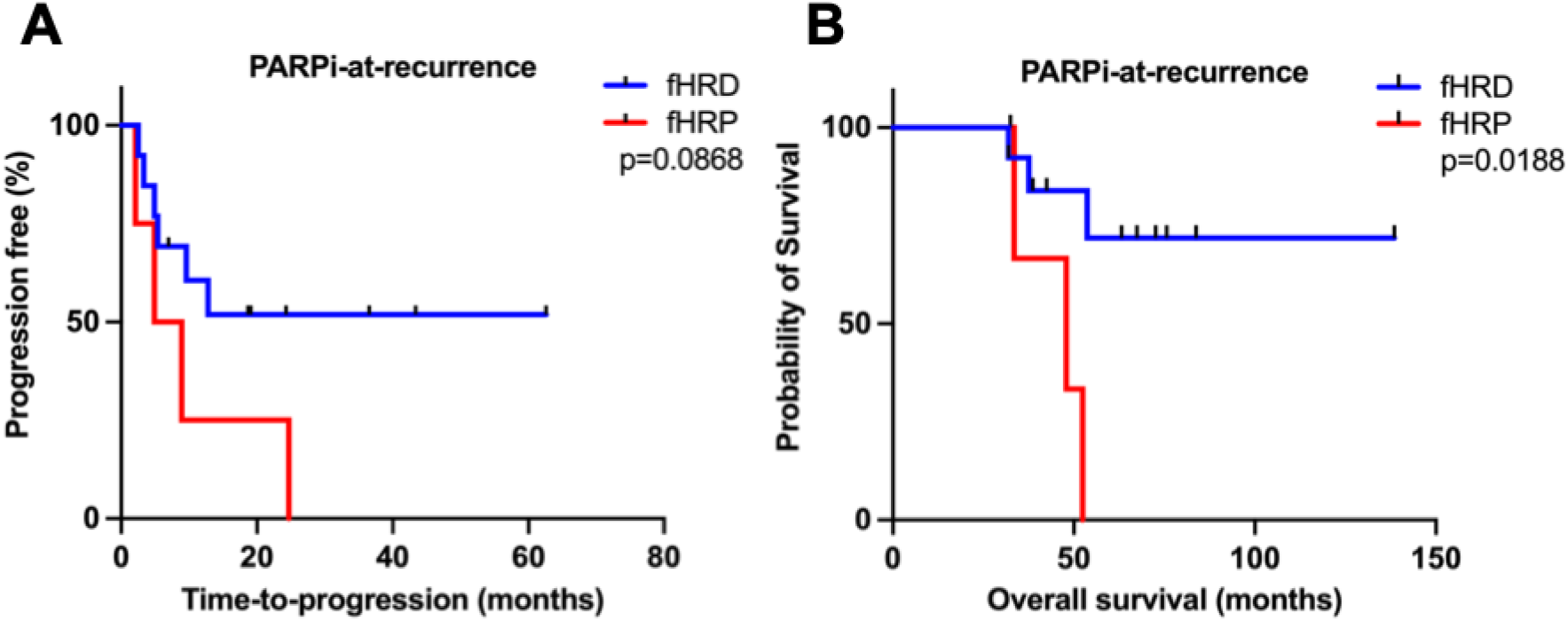
fHRD status indicates PARPi treatment sensitivity at recurrence. **A**. In Kaplan-Meier survival analysis time-to-progression from PARPi treatment start did not significantly differ between fHRD and fHRP patients (Log-rank, Mantel-Cox test). **B**. OS of patients who had received PARPi treatment at recurrence was significantly longer in the fHRD group than in the fHRP group (Log-rank, Mantel-Cox test).

### Comparison between different HRD tests

In addition to the functional HR test, we also had three genomics-based HRD estimates – SBS3, ID6 and ovaHRDScar – available for most of the specimens sampled at Turku University Hospital. In univariate hazard ratio analyses, in addition to fHR status (hazard ratio: 0.255, p<0.0001), ovaHRDScar was found to significantly correlate with longer PFI (hazard ratio: 0.3243, p=0.0089, **Fig. 5A**). For HRD groups, fHRD, SBS3+, ovaHRDScar+ or ID6+, mean platinum-free intervals were 17.5, 14.85, 18.1 and 15.2 months, respectively. For HRP groups, fHRP, SBS3-, ovaHRDScar- and ID6-, mean platinum-free intervals were 5.2, 7.55, 6.1 and 9 months, respectively.

**Figure 5.**
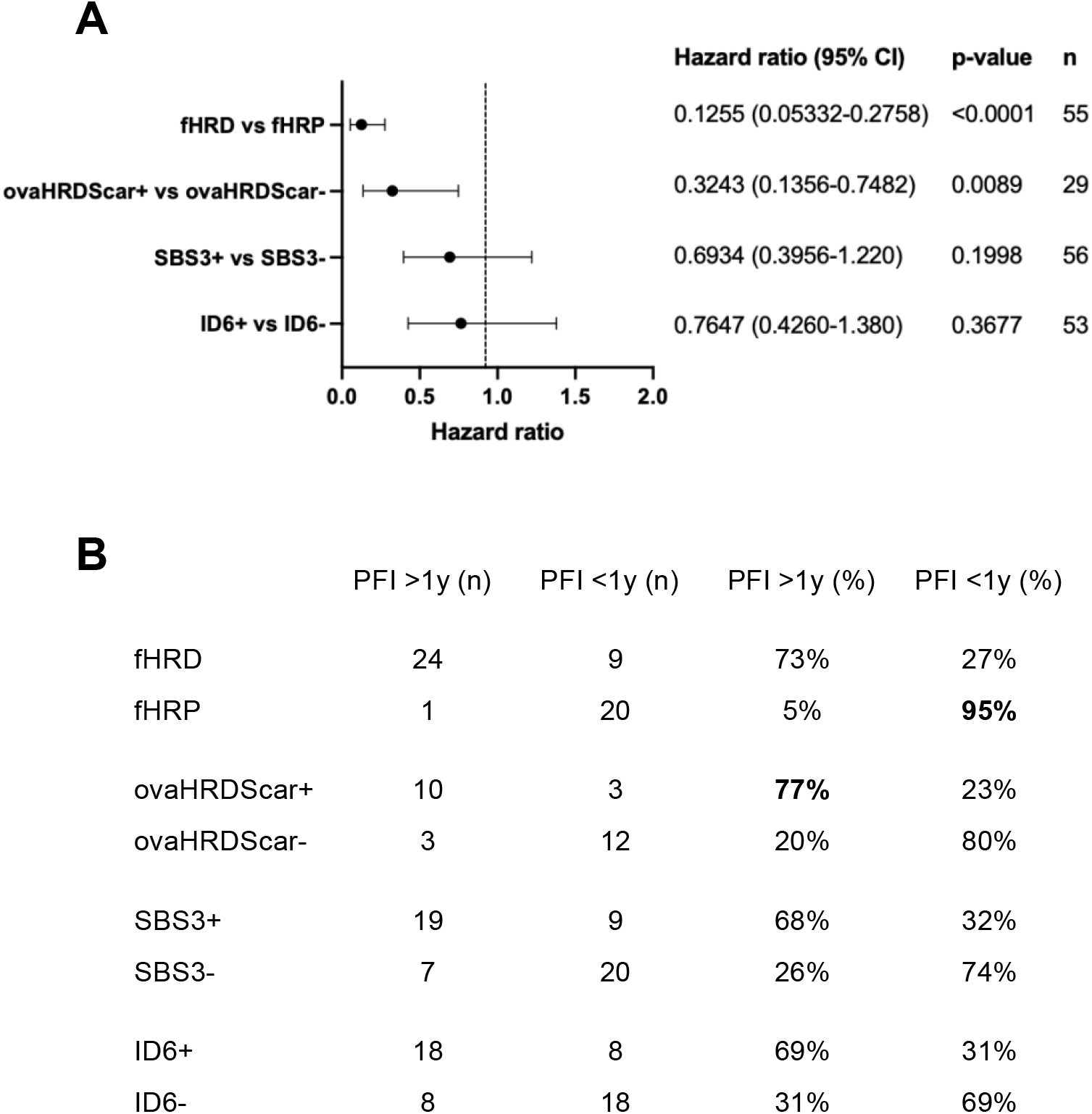
**A**. Univariate hazards ratio analysis with the different HRD tests available. Only fHR and ovaHRDScar tests produce significant results (Cox proportional hazards regression). **B**. Sensitivity and specificity of the different HRD tests. The ovaHRDScar has the best sensitivity in detecting HR deficiency (>1y PFI) and fHR test has the best specificity in detecting HR proficiency (<1y PFI).

We also compared the sensitivity and specificity of the different HRD tests (**Fig. 5B**). The ovaHRDScar test identified patients with durable response to the first-line platinum therapy as HRD (PFI >1 year) with the best sensitivity (77% versus fHRD 73%, SBS3+ 68% and ID6 69%), while fHR test identified patients with <1-year PFI as HRP with the best specificity (95% versus ovaHRDScar-80%, SBS3-74% and ID6-69%).

## Discussion

HGSCs are often defective in homologous recombination repair of DNA double-strand breaks, and patients with such tumors have better response to platinum and PARPi therapy. PARPi treatment has revolutionized HGSC treatment, but a “PARPi for all” strategy is not viable in the long term, due to high cost and appreciable side effects (Wethington et al., 2021). Not all patients respond to platinum-based therapy or PARPi and should be channeled for other drug treatments. Thus, it is critical to identify patients with HRD tumors, to target the right therapy to the right patients. Assaying RAD51 function, rather than HRD-associated genomics, is an attractive approach, as it measures current, not historical, HR capacity of tumor cells and is mutation-agnostic. FFPE-based RAD51 assays have previously been described in breast cancer samples (Castroviejo-Bermejo et al., 2018; Cruz et al., 2018; Pellegrino et al., 2022), epithelial ovarian cancer (Hoppe et al., 2021) and ovarian/endometrial cancer sample cohorts (van Wijk et al., 2021), but never for advanced HGSC specifically. No published studies exist on functional HR testing from IDS samples, which constitute an appreciable fraction of available HGSC specimens. Moreover, very limited data exist (Hoppe et al., 2021; Tumiati et al., 2018; van Wijk et al., 2020) on the predictive value of functional HR scores for *in vivo* therapy response in patients with advanced HGSC. Here, we show that RAD51 quantification in FFPE sections (fHR test) of unselected HGSC patients can identify functional HRD beyond BRCAmut tumors and significantly predict platinum response and other key clinical outcomes. Importantly, we also established a novel fHRD scoring protocol for diagnostic IDS FFPE blocks that predicts clinical response to HGSC therapy. Further, the fHR test shows promise for identifying patients who benefit most from PARPi.

To our knowledge, this is the first demonstration that functional HR status from IDS specimens, that is samples that have been exposed *in vivo* to a full course of standard-of-care NACT treatment, significantly predicts therapy response, as measured by different clinical outcomes. Previously, the Vreeswijk group determined HR capacity using an *ex vivo* irradiation protocol in a cohort consisting of PDS and IDS specimens, but no significant correlation with PFI or OS was found, nor were results from IDS samples reported separately (van Wijk et al., 2020). Our results demonstrate that for NACT-treated IDS samples the proposed 30% cut-off value for fHRD significantly predicts two key clinical outcomes, PFI and OS. Its robustness should be tested in larger, independent cohorts of NACT-treated patients.

The results reported here are, to our knowledge, the first to show the predictive power of functional HRD – as measured from routine HGSC FFPE specimens – for real-world, clinical *in vivo* platinum and PARPi responses. An added benefit of our analysis method is that it does not include counting individual RAD51 foci per nucleus, but instead defines each nucleus as RAD51-positive or - negative, making the assay more robust and circumventing the need for confocal microscopy. Another major strength of the fHR assay is that it can be performed on tumor samples with only a few proliferating tumor cells. This is critical as NACT-treated patients who have good platinum response often have very few proliferating tumor cells left when they undergo IDS surgery. These patients are precisely those most likely to derive significant benefit from PARPi, but currently would be unscorable with genomics-based clinical HRD tests, as these assays require relatively high tumor percentage.

Naturally, the fHR assay is not without limitations: it can identify only those functionally HRD samples with impaired RAD51 loading. Also, a sample must have sufficient endogenous DNA damage in G2/S phase cells (>10%) to generate a reliable fHR score. Of our HGSC samples, however, only three (2.1%) were excluded for too-low DNA damage in G2/S phase. Thus, it seems that the vast majority of routine HGSC samples have enough endogenous DNA damage to obtain a fHR score.

Two out of 16 BRCAmut tumors were scored as fHRP (EOC378 and EOC105) and perhaps coincidentally, both cases had somatic *BRCA2* mutations and both were found, by WGS, to have reversion mutation in their relapse sample. Our functional HR test, performed on the primary tumor sample, already categorized these patient as fHRP, perhaps due to the presence of the HRP (BRCA2-revertant) subclone. PFI of EOC378 was 12.6 months indicating platinum sensitivity and HR deficiency, whereas EOC105 had a PFI of 6.5 months indicating a more platinum resistant disease and HR proficiency. It should be noted, that not all BRCAmut cases behave clinically as expected, for instance, in the SOLO1 trial, approximately 10% of germline BRCAmut patients progressed within 12 months despite PARPi maintenance therapy, and 50% of germline BRCAmut patients in the placebo group failed to show a durable (>12 month) platinum response (Moore et al., 2018). As such, one should be cautious about BRCAmut status as the ground truth for HRD. Ultimately, the ground truth that is most relevant in the context of HRD testing is real-world clinical response to platinum and/or PARPi.

All HRD tests are plagued by some degree of false negative and false positive HRD calls. For example, genomically HRD patients – as defined by BRCAmut status and/or Myriad GIS – were not significantly enriched in PARPi long-term responders in the recurrent setting (Lheureux et al., 2017). Arguably, the most pertinent task is to better identify the BRCAwt HRD patient population, given that many PARPi long- and intermediate-term responders are BRCAwt (Swisher et al., 2021a). To this end, our results demonstrate that the fHR test significantly predicts long-term platinum response (a reasonable proxy for PARPi response), even when BRCAmut cases are removed from analyses.

Although the number of PARPi-treated patients in our cohort was limited, this report is, to our knowledge, the first to correlate functional HR status with *in vivo* PARPi response in HGSC patients. This retrospective analysis indicates that the fHR test may be particularly useful for identifying patients who fail to respond to PARPi (those with fHRP tumors), at least in the recurrent setting. Further strengthening the idea that fHRP status reports on HRP-like clinical behavior is the fact that fHRP patients displayed significantly worse platinum responses, both to first- and second-line treatment. This finding is in line with a recent report that high RAD51 nuclear expression scores associate with worse platinum response in ovarian cancer (Hoppe et al., 2021). fHRP patients could in the future be channeled for alternative treatments, at least in the recurrent setting, as they are not likely to respond to platinum-based chemotherapy or to PARPi.

Our findings provide a strong motivation for including fHR testing into prospective PARPi clinical trials, and/or retrospectively analyzing archival FFPE blocks from PARPi trials. The fHR test, as reported here, can be performed on routine FFPE samples, and has several additional practical advantages: it does not matter at which type of surgery the specimen is acquired (laparoscopic, PDS and IDS), nor from which anatomical location. It should be emphasized, however, that short time-to-fixation (<2 hours) is needed for successful fHR testing, potentially limiting the utility of some archival FFPE material.

In summary, fHRD testing shows great promise as a universal method for stratifying patients with advanced HGSC as platinum- and PARPi-sensitive. The fHR score significantly predicts clinical response, both PFI and OS, and is indicative of PARPi sensitivity. We propose it should be incorporated into the HRD testing toolkit to support clinical decision making.

## Data Availability

All data produced in the present study are available upon reasonable request to the authors.

## Acknowledgements

We thank Sonja Koopal, Taina Turunen, Alisha GM, Saundarya Shah, Virpi Asikainen, Ella Lintunen, Victoria Kuusinen, Nina Halme, Tiina Vartiala and Maija Vääriskoski for excellent technical assistance. We also wish to thank the staff at the tissue preparation and histochemistry unit of the Anatomy department at University of Helsinki and the Finnish Institute of Molecular Medicine (FIMM) Digital Microscopy and Molecular Pathology Unit at Helsinki Institute of Life Science (HiLIFE) and University of Helsinki. Images were generated using 3DHISTECH Pannoramic 250 FLASH II digital slide scanner at the Genome Biology Unit supported by HiLIFE and the Faculty of Medicine, University of Helsinki, and Biocenter Finland.

