## Supplemental Data for "Functional homologous recombination assay on FFPE specimens of advanced high-grade serous ovarian cancer predicts clinical outcomes"

**S1**

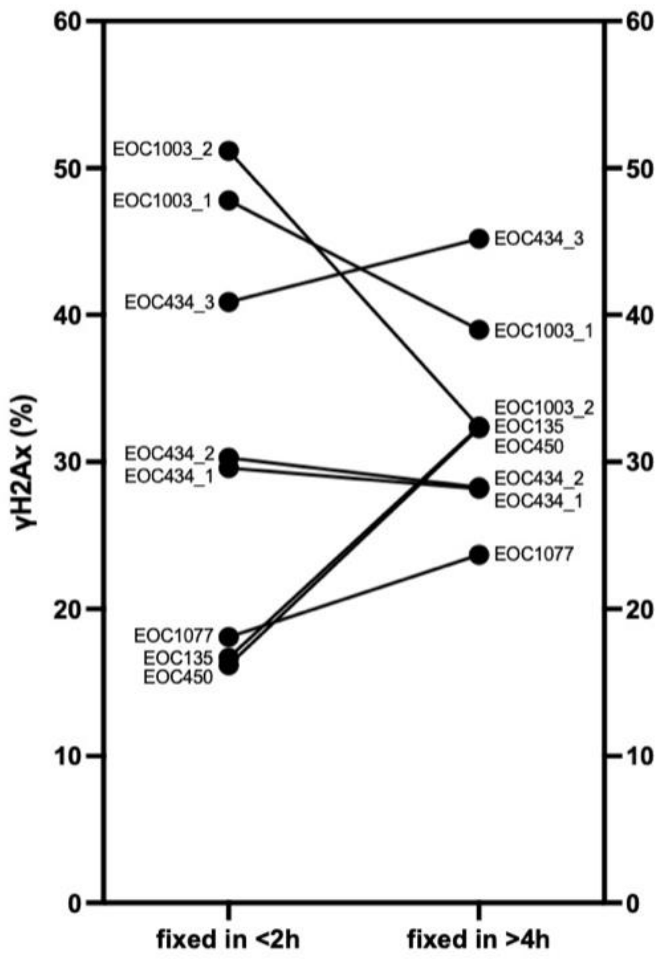


***Supplementary figure 1.*** *Amount of DNA damage, marked by γH2Ax, in samples fixed in <2h and >4h after resection. Large variation in the amount of DNA damage (γH2Ax) can be seen between samples that have been fixed in <2 hours and samples that have been fixed after >4 hours from resection.*

## S2

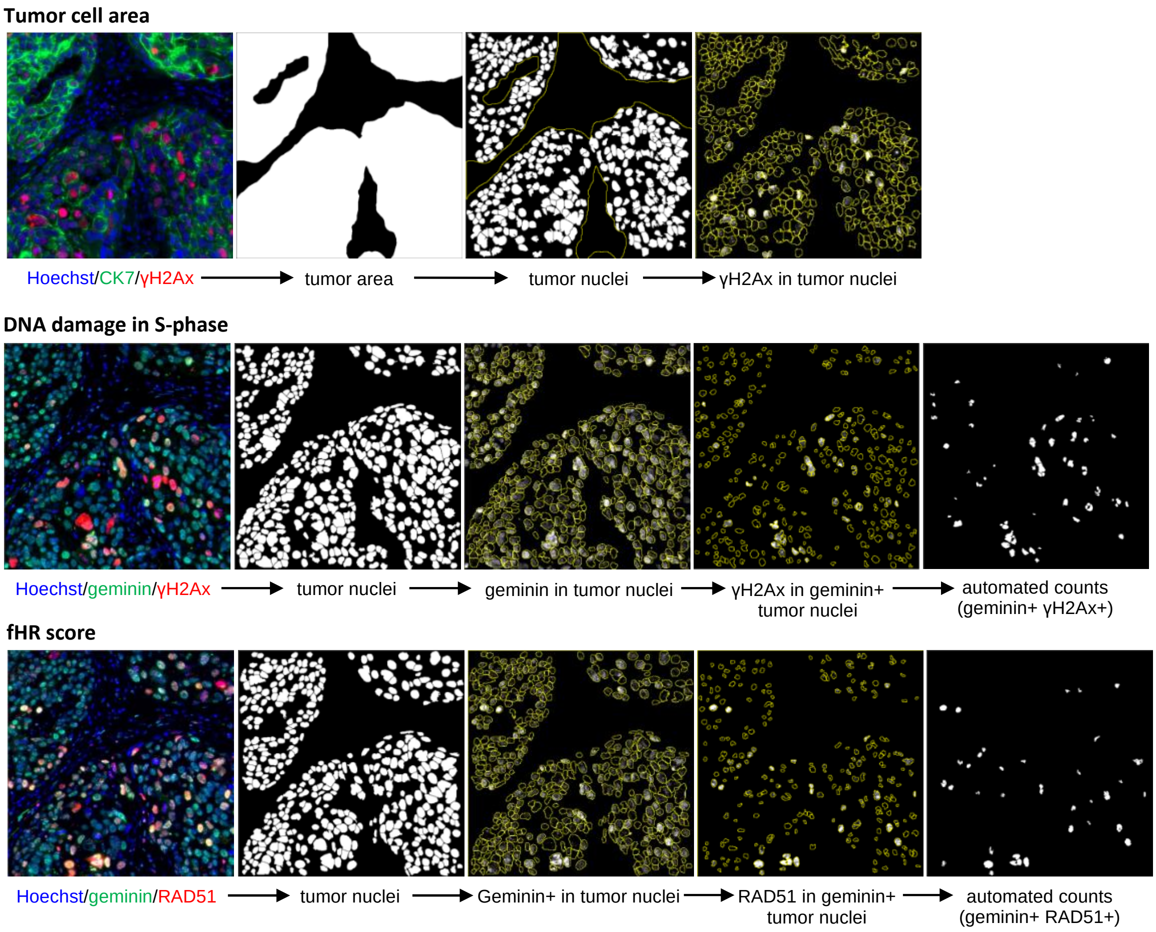


***Supplementary figure 2.*** *Image analysis workflow in ImageJ. From CK7-γH2Ax double stained section, the tumor cell areas and areas with DNA damage are identified. From the geminin-γH2Ax double stained section, the amount of DNA in S/G2 phase cells is quantified. Functional HR score is calculated as the percentage of RAD51+geminin+ nuclei out of all geminin+ nuclei form the geminin-RAD51 double stained section.*

**S3**

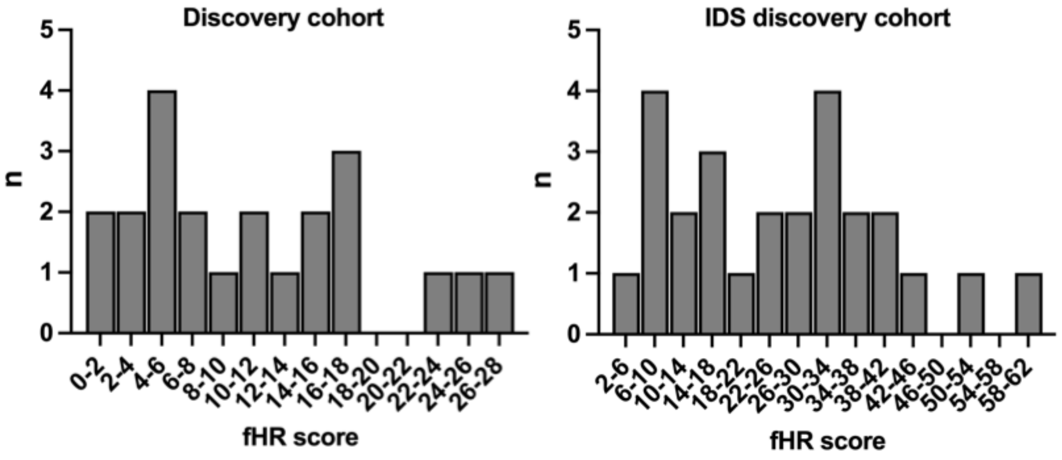


***Supplementary figure 3.*** *Distributions of fHR scores in chemo-naïve and IDS discovery cohorts.*

## S4

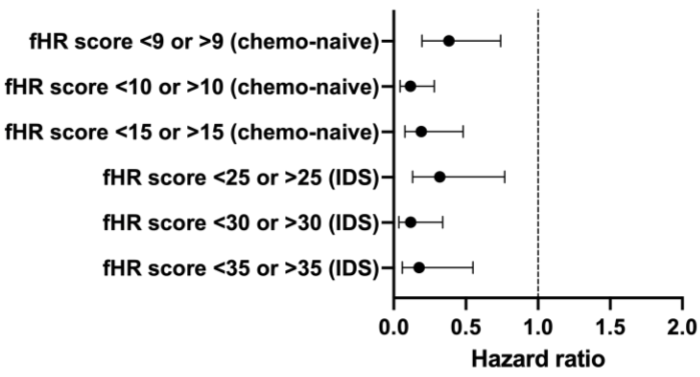


| Hazard ratio (95% CI) | p-value |
|---|---|
| 0.3828 (0.1964-0.7405) | 0.0043 |
| 0.1162 (0.04424-0.2806) | <0.0001 |
| 0.1904 (0.07723-0.4814) | 0.0003 |
| 0.3196 (0.1302-0.7690) | 0.0108 |
| 0.1171 (0.03554-0.3388) | 0.0001 |
| 0.1757 (0.05934-0.5486) | 0.0017 |

***Supplementary figure 4.*** *Hazard ratio analysis with different cut-off values for fHR score. All tested cut-off values produced a significant result, but 10 for chemo-naive and 30 for chemo-treated had narrower 95% confidence interval (univariate Cox proportional hazards regression).*

**S5**

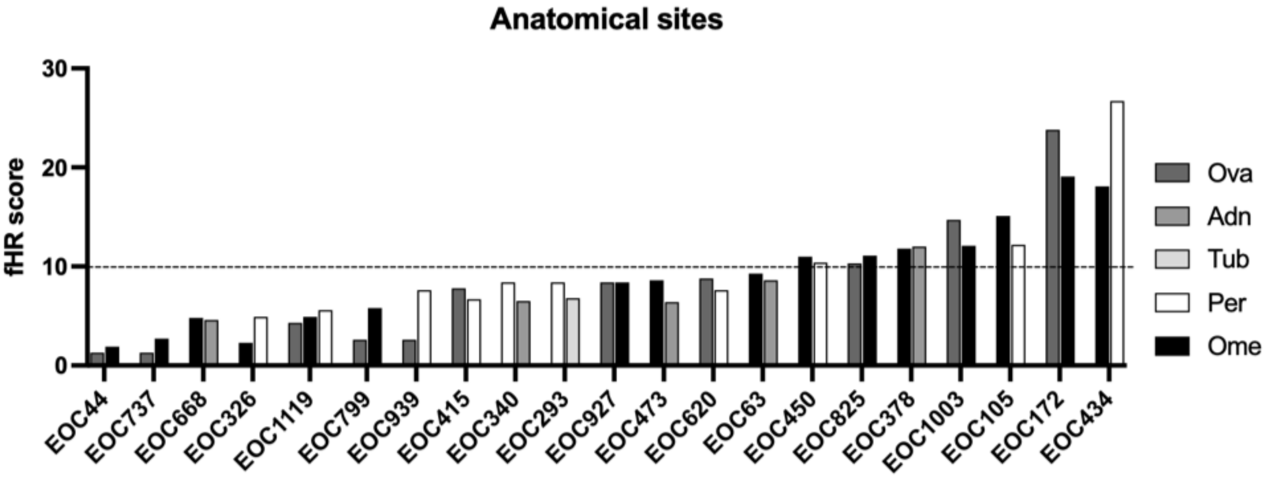


**Supplementary figure 5.** Comparison between fHR scores from chemonaive samples obtained from different anatomical locations of individual patients. Although the numerical value of fHR score differs between sample locations, the fHR status (fHRD vs fHRP) stays the same. Abbreviations: Ova, ovary; Adn, adnex; Tub, fallopian tube; Per, peritoneum; Ome, omentum.

S6

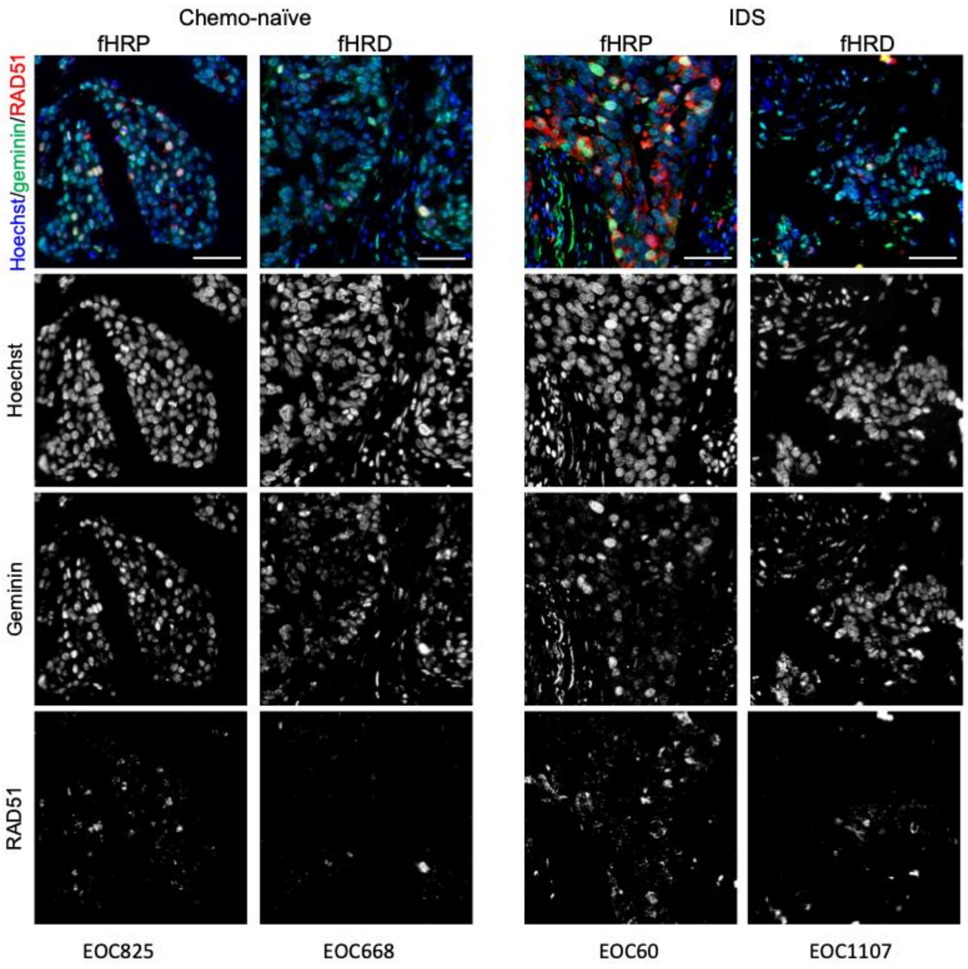


***Supplementary figure 6.*** *Example images of staining patterns in chemo-naive and NACT-treated (IDS) geminin-RAD51 double stained samples. Scale bar 50μm.*

**S7**

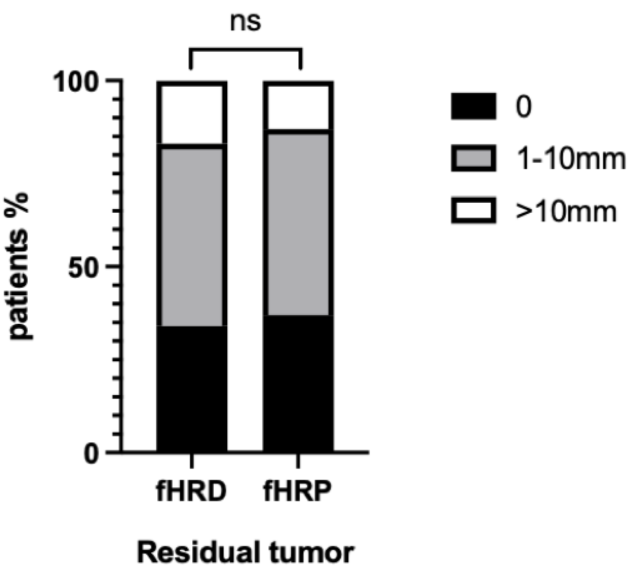


***Supplementary figure 7.*** *Comparison between the success of cytoreduction in fHRD and fHRP groups. No difference between the fHRD and fHRP groups was found in the success of cytoreduction (Chi-square test).*

**S8**

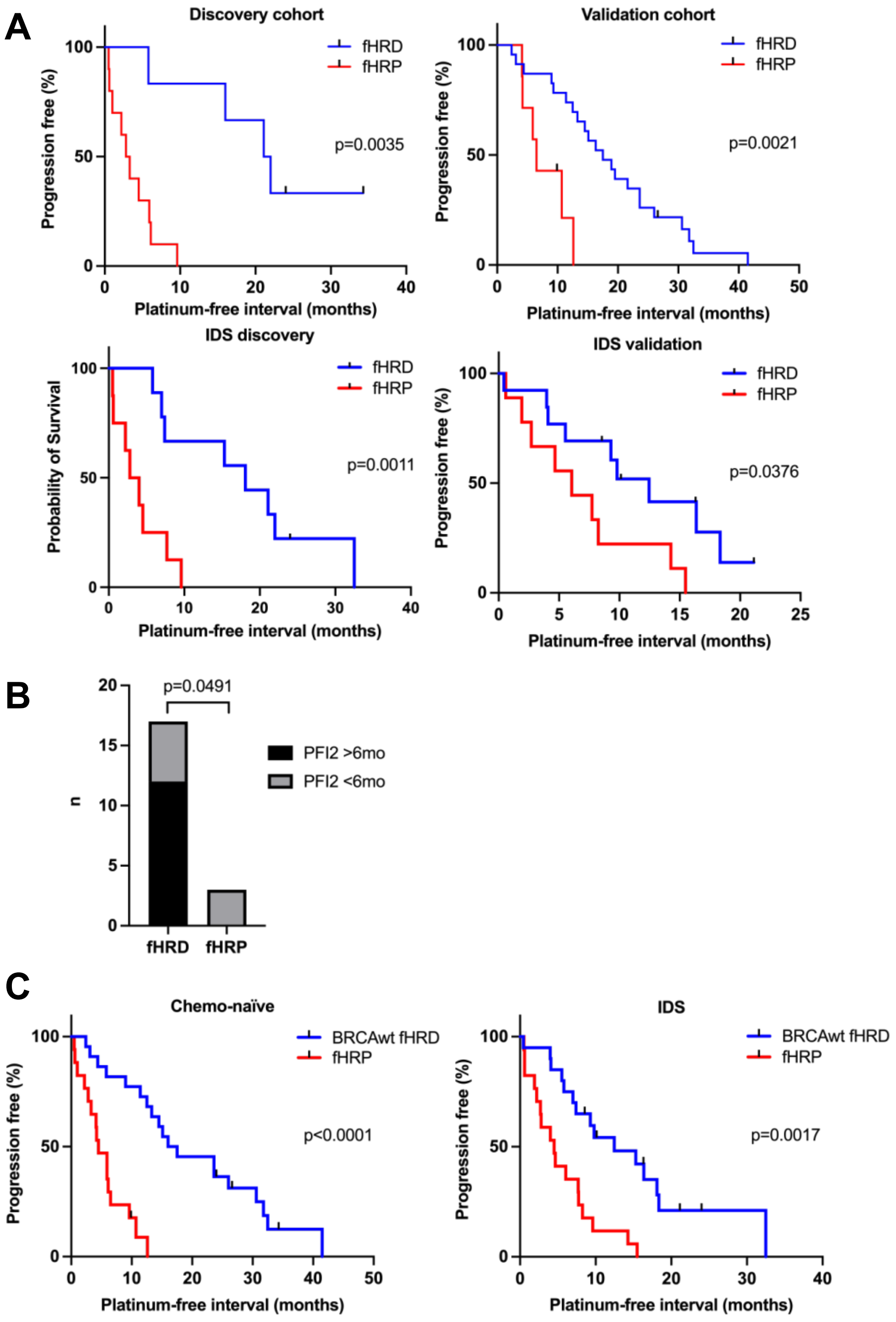


***Supplementary figure 8.*** *Platinum-free interval and overall survival in the different cohorts.* ***A.*** *PFI shown separately in discovery and validation cohorts of chemo-naïve and IDS samples (Log-rank, Mantel-Cox test).* ***B.*** *Response to second-line platinum treatment in fHRD (n=17) and fHRP (n=3) groups (Fisher's exact).* ***C.*** *PFI in chemo-naive and chemo-treated (IDS) cohorts without BRCAmut fHRD patients (Log-rank, Mantel-Cox test).*
