## Supplemental Table 1 for "Functional homologous recombination assay on FFPE specimens of advanced high-grade serous ovarian cancer predicts clinical outcomes"

| Patient ID | HRR mutation | somatic/germline | Somatic HRR mutation in relapse only |
|---|---|---|---|
| EOC955 | BRCA1:NM_007294:c.3626delT:p.L1209*:TA/T | germline | ND |
| EOC939 | BRCA1:NM_007294:c.5444G>A:p.W1815X | somatic | ND |
| EOC933 | BRCA2:NM_000059:c.1338delG:p.L446Ffs*14 | somatic | ND |
| EOC927 | BRCA1:NM_007294:c.5266dupC:p.Q1756Pfs*74:T/TG | germline | ND |
| EOC890 | | | |
| EOC87 | | | ND |
| EOC868 | BRCA2:NM_000059:c.8960delT:p.L2987Rfs*14 | somatic | |
| EOC855 | | | ND |
| EOC825 | | | ND |
| EOC809 | | | |
| EOC799 | | | ND |
| EOC763 | BRCA1:BRCA1:NM_007294:del:5_exon_deletion | somatic | ND |
| EOC752 | | | |
| EOC742 | | | |
| EOC737 | BRCA1:NM_007294:c.1204G>T:p.E402X | somatic | ND |
| EOC733 | | | ND |
| EOC691 | BRCA1:NM_007294:c.604C>T:p.Q202X; FANCM:NM_000136:c.81_82insTGGCATTCTTCCTGAAAAAGATTTCT CAGTTGAAGAATTTGCCTCTCTTCAAATATGCCATTTTGTA:p.G28Wfs*5 | somatic | ND |
| EOC668 | | | ND |
| EOC664 | ND | | ND |
| EOC649 | | | ND |
| EOC63 | BRCA1:NM_007294:c.3626delT:p.L1209*:TA/T | germline | ND |
| EOC620 | | | ND |
| EOC600 | BRCA2:NM_000059:c.9118-2A>G:NA:A/G | germline | ND |
| EOC60 | ND | | ND |
| EOC568 | | | ND |
| EOC561 | CDK12:NM_016507:c.365T>G:p.L122X | somatic | ND |
| EOC540 | | | ND |
| EOC498 | | | ND |
| EOC495 | | | |
| EOC473 | | | ND |
| EOC450 | RAD51C:NM_058216:exon1:c.90delG:p.F32Sfs*8 | germline | PALB2:NM_024675:c.1501delA:p.R501Gfs*60 |
| EOC44 | | | ND |
| EOC434 | | | |
| EOC423 | CDK12:NM_016507:c.2644C>T:p.R882W | somatic | ND |
| EOC415 | | | |
| EOC412 | RAD51C:NM_058216:c.90delG:p.F32Sfs*8:CG/C | germline | ND |
| EOC389 | | | ND |
| EOC388 | | | ND |
| EOC382 | | | ND |
| EOC378 | BRCA2:NM_000059:c.2978_2982del:p.A994Tfs*13:TGGGCA/T | germline | BRCA2:NM_000059:c.2987_2993del:p.L997Qfs*44 |
| EOC376 | | | ND |
| EOC372 | | | ND |
| EOC341 | ND | | ND |
| EOC340 | | | ND |
| EOC336 | BRCA1:NM_007294:exon10:c.3606_3652del:p.Y1202X | somatic | ND |
| EOC326 | | | ND |
| EOC3 | FANCC:NM_000136:exon8:c.798_799insGA:p.N267Efs*4 | germline | ND |
| EOC295 | | | |
| EOC293 | CDK12:NM_016507:c.1745C>G:p.S582X | somatic | ND |
| EOC286 | | | ND |
| EOC26 | | | |
| EOC256 | | | ND |
| EOC227 | FANCI:NM_001113378:exon27:c.2957_2969del:p.V986Afs*39 | germline | |
| EOC218 | | | ND |
| EOC207 | | | ND |
| EOC175 | | | ND |
| EOC172 | | | |
| EOC160 | | | |
| EOC151 | | | ND |
| EOC135 | | | |
| EOC133 | BRCA2:NM_000059:c.8331+2T>C:NA:T/C | germline | ND |
| EOC126 | | | ND |
| EOC1133 | BRCA2:NM_000059:exon18:c.8023A>G:p.I2675V | somatic | ND |
| EOC1129 | | | ND |
| EOC1120 | | | |
| EOC1119 | | | ND |
| EOC1107 | | | ND |
| EOC1077 | BRCA1:NM_007294:exon10:c.1082_1092del:p.S361* | germline | ND |
| EOC105 | BRCA2:NM_000059:c.6591_6592del:p.E2198Nfs*4 | somatic | BRCA2: NM_000059:g.32339567-32341857del |
| EOC103 | | | ND |
| EOC1023 | ND | | ND |
| EOC1005 | | | ND |
| EOC1003 | | | ND |
| EOC1002 | FANCM:NM_020937:exon20:c.5101C>T:p.Q1701X | germline | ND |
| C985 | | | ND |
| C972 | | | ND |
| C956 | | | ND |
| C946 | BRCA1 (from clinical testing) | | ND |
| C917 | | | ND |
| C911 | | | ND |

| | | | |
|---|---|---|---|
| C893 | | | ND |
| C799 | | | ND |
| C790 | | | ND |
| C686 | | | ND |
| C569 | BRCA1 (from clinical testing) | | ND |
| C546 | | | ND |
| C538 | BRCA2 (non-pathogenic, from clinical testing) | | ND |
| C452 | | | ND |
| C433 | | | ND |
| C382 | | | ND |
| C369 | | | ND |
| C344 | | | ND |
| C331 | | | ND |
| C137 | | | ND |
| C129 | | | ND |
| C124 | | | ND |
| C091 | | | ND |
| C079 | | | ND |
| C063 | | | ND |
